# Gut microbiota predict *Enterococcus expansion* but not Vancomycin-resistant *Enterococcus* acquisition

**DOI:** 10.1101/2020.05.26.20114181

**Authors:** Rishi Chanderraj, Christopher A. Brown, Kevin Hinkle, Nicole Falkowski, Piyush Ranjan, Robert P. Dickson, Robert J. Woods

## Abstract

**BACKGROUND:** Vancomycin-resistant Enterococcus (VRE) is a leading cause of hospital-acquired infections, and continues to spread despite widespread implementation of pathogen-targeted control guidelines. Commensal gut microbiota provide colonization resistance to VRE, but the role of gut microbiota in VRE acquisition in at-risk patients is unknown.

**METHODS:** We performed a case-control study of gut microbiota in hospitalized patients who did (*cases*) and did not (*controls*) acquire VRE. We matched case subjects to control subjects by known risk factors and “time at risk,” defined as the time elapsed between admission until positive VRE screen. We characterized gut bacterial communities using 16S rRNA gene amplicon sequencing of rectal swab specimens.

**RESULTS:** We analyzed 236 samples from 59 matched case-control pairs. At baseline, case and control subjects did not differ in gut microbiota when measured by community diversity (p=0.33) or composition (p=0.30). After hospitalization, gut communities of cases and controls differed only in the abundance of the *Enterococcus* containing operational taxonomic unit (OTU), with the gut microbiota of case subjects having more of this OTU than time-matched control subjects (p=0.01). Otherwise, case and control communities after the time at risk did not differ in diversity (p=.33) or community structure (p=0.12). Among patients who became VRE colonized, those having the *Blautia* containing OTU on admission had lower *Enterococcus* relative abundance once colonized (p =0.004).

**CONCLUSIONS:** The 16S profile of the gut microbiome does not predict VRE acquisition in hospitalized patients, likely due to rapid and profound microbiota change. The gut microbiome does not predict VRE *acquisition*, but may be associated with *Enterococcus expansion*, suggesting that these should be considered as two distinct processes.

**Summary:** 16S profile of the gut microbiome does not predict VRE acquisition in hospitalized patients, likely due to the rapid and profound microbiota change in this) opulation. VRE expansion and VRE acquisition may be two distinct processes.

## INTRODUCTION

Vancomycin-resistant Enterococcus (VRE) is a highly antibiotic-resistant bacteria, is a leading cause of healthcare-associated infections, and is classified as a serious public health threat by the Centers for Disease Prevention and Control[4,5]. Colonization with VRE precedes infection[6,7], and molecular epidemiologic analyses show patient-to-patient hospital transmission is the primary means of spread[8]. Preventing transmission between hospitalized patients is a significant challenge, and despite the widespread application of pathogen-targeted control measures[9], VRE remains endemic in many hospitals[4,5].

Both indirect human evidence and animal experimentation demonstrate that gut microbiota prevent VRE colonization when a patient is exposed, a phenomenon termed “colonization resistance” [2,10,11]. Colonization resistance may entail competition for resources, secretion of bactericidal factors[3,12], and indirect stimulation of host immune defense mechanisms that target VRE[13,14]. Though colonization resistance plays a crucial role in suppressing VRE expansion and preventing VRE infection[1,15], to date, no study has evaluated whether variation in intestinal microbiota can explain variation in VRE *acquisition* among at-risk patients.

To address this gap in our understanding of VRE transmission, we investigated whether the gut microbiome of at-risk patients predicts VRE colonization in a hospitalized patient population. We hypothesized that if the gut microbiome can confer colonization resistance for VRE acquisition, variation in baseline microbiota would explain variation in patient susceptibility to VRE acquisition. To test this hypothesis, we designed a case-control study using 16S rRNA gene amplicon sequencing of rectal swabs acquired from hospitalized patients.

## METHODS

### Study setting and design

We previously conducted a retrospective case-control study of clinical risk factors for VRE acquisition among patients who did (cases) and did not (controls) acquire VRE during their admissions at the University of Michigan Healthcare System from January 2013 until June 2016[16]. We studied gut microbiome communities in 236 rectal swab samples from 59 matched pairs of case and control subjects from patients admitted to the University of Michigan Hospital in 2016. 64 out of 118 subjects in this study (54%) were a part of our previous clinical risk factor analysis. The remaining subjects were admitted from June to December 2016 (outside the timeframe of the previous study by six months). The University of Michigan Healthcare system consists of ~1000 inpatient beds and serves as a tertiary referral hospital for southeastern Michigan. The institutional review board at the University of Michigan approved the study before its initiation.

The infection control practice throughout the study period was to perform routine surveillance for VRE on eight adult units, including intensive care units, the hematology and oncology ward, and the bone marrow transplant ward. All patients were routinely screened on admission and weekly thereafter with rectal swabs that were tested by Bio-Rad VRESelect™ chromogenic medium to detect VRE. Cases were defined as subjects with an initial negative swab followed by a positive swab when evaluated by this selective culture. We further identified the “time at risk” for each case patient, here defined as the time elapsed between admission and positive VRE screen. We matched each case subject to a control subject with an initial negative swab followed by repeat negative swab within the same time at risk (+/-5%). An additional matching factor was the unit from which the first positive VRE was recovered for cases or the matched swab after the time at risk for controls.

### Bacterial DNA isolation

Genomic DNA was extracted from rectal swabs re-suspended in 360 μl ATL buffer (Qiagen DNeasy Blood & Tissue kit) and homogenized in fecal DNA bead tubes using a modified protocol previously demonstrated to isolate bacterial DNA[17,18]. Sterile laboratory water and AE buffer used in DNA isolation were collected and analyzed as potential sources of contamination. ZymoBIOMICS Microbial Community DNA Standard (Zymo Research cat# D6306) was sequenced for error analysis.

### 16s rRNA gene sequencing

The V4 region of the 16s rRNA gene was amplified using published primers and the dual-indexing sequencing strategy developed previously [17]. Sequencing was performed using the Illumina MiSeq platform (San Diego, CA), using a MiSeq Reagent Kit V2 (500 cycles), according to the manufacturer’s instructions with modifications found in the standard operating procedure of the laboratory of Dr. Patick Schloss [17,19]. Accuprime High Fidelity Taq was used in place of Accuprime Pfx SuperMix[20]. Primary PCR cycling conditions were 95°C for two minutes, followed by 20 cycles of touchdown PCR (95°C 20 seconds, 60°C 20 seconds and decreasing 0.3 degrees each cycle, 72°C for 5 minutes), then 20 cycles of standard PCR (95°C for 20 seconds, 55°C for 15 seconds, and 72°C for 5 minutes), and finished with 72°C for 10 minutes.

### Statistical analyses

Sequence data were processed and analyzed using the software mothur v.1.43.0[21] according to the standard operating procedure for MiSeq sequence data using a minimum sequence length of 250 base pairs[17,22]. To summarize, the SILVA rRNA database[23] was used as a reference for sequence alignment and taxonomic classification. K-mer searching with 8mers was used to assign raw sequences to their closest matching template in the reference database, and pairwise alignment was performed with the Needleman-Wunsch[24] and NAST algorithms[25]. A k-mer based Naïve Bayesian classifier[26] was used to assign sequences to their correct taxonomy with a bootstrap confidence score threshold of 80. Pairwise distances between aligned sequences were calculated with the method employed by Sogin et al. [27], where pairwise distance equals mismatches, including indels, divided by sequence length. A distance matrix was passed to the OptiCLUST clustering algorithm[28] to cluster sequences into ‘Operational Taxonomic Units’ (OTUs) by maximizing the Matthews Correlation Coefficient with a dissimilarity threshold of 3%[29].

After clustering and classification of raw sequencing data, we evaluated differences in community structure with Permutational multivariate analysis of variance (PERMANOVA) in the *vegan* package (v 2.0-4)[30] in R (v 3.6.4) [31]. We performed resampling of multiple generalized linear models with the *mvabund[32]* package in R to look for individual OTU differences between communities. We set a significance threshold of 0.01 after adjusting for multiple comparisons using a stepdown resampling procedure to reduce the type I error rate[33]. We confirmed individual OTU differences with random forest classification and regression models built with the *ranger* package in R (v 0.11.2) [34]. We used the *caret* (v 6.0-84)[35] package in R for cross-validation and to optimize the hyperparameters of the number of decision trees in the model and the number of features considered by each tree when splitting a node. We corrected for feature importance bias in random forest models with a permutation importance (PIMP) heuristic developed by Altmann et al. [36]. All OTUs were included in diversity and abundance analyses. We performed bivariate analysis with conditional logistic regression using the *survival* (v 3.1-8) package in R[37,38]. Differences in means of final *Enterococcus* abundance in *cases only* were compared with the non-parametric Mann-Whitney U-Test. We used the *vegan* package in R to calculate both the average species diversity in an individual rectal swab (Shannon Diversity) and the change in community structure between initial swab and second swab for each subject (Jaccard Distance). We used Spearman’s rank correlation coefficient to determine if Jaccard distance was significantly correlated with the time between swabs, and used non-linear least squares regression to fit a model of Jaccard Distance over time for cases and controls.

### Adequacy of Sequencing

We performed 16S rRNA gene amplicon sequencing on 236 rectal swab specimens and 15 negative control specimens, which identified 1,188 unique operational taxonomic units (OTUs, genus-level bacterial taxa) at a dissimilarity threshold of 3%. After bioinformatics processing, the mean number of reads per sample was 71,484±2,684. No specimens were excluded from the analysis. Rectal swab specimens had clear differences in community structure compared to control specimens, which was confirmed as statistically significant using multiple methods of hypothesis testing [mvabund and PERMANOVA (adonis), p < 0.01 both] (**Supplemental figure 1**). Sequences generated from negative control specimens were dominated by a single *Pseudomonas-classified* OTU (OTU001). This OTU was included in all reported analyses, though the exclusion of this OTU did not affect any of the reported results.

### Data availability

Sequences are available via the NCBI Sequence Read Archive (accession number PRJNA633879). OTU tables, taxonomy classification tables, and metadata tables are available at https://github.com/rishichanderraj/Microbiota_Predictors_VRE_Acquisition.

## RESULTS

### Study population and medication exposures

We studied gut microbiome communities in 236 rectal swab samples from 59 matched pairs of case and control subjects (**Table 1**). Cases and controls did not differ in demographics (age, sex, ethnicity) nor in the relative frequency of common comorbidities (e.g. immunosuppression, malignancy, or gastrointestinal disease). Antibiotic use was widespread among all subjects and was nearly equal across groups (**Table 2**). Vancomycin, cefepime, metronidazole, and piperacillin-tazobactam were the most commonly used antibiotics in the study population. Cases and controls did not differ significantly in their exposure to any specific antibiotics prior to initial sampling. More cases received proton pump inhibitors prior to initial sampling (P = 0.04). During time-at-risk (between initial and subsequent sampling), case and controls did not differ in their exposure to antibiotics or proton pump inhibitors.

**Table 1.** Demographics and Comorbidities of Matched Cohorts

|  | *Controls*<br>n = 59 | *Cases*<br>n = 59 | *P*-value |
| --- | --- | --- | --- |
| **Demographics** |  |  |  |
| Age (mean ± SE) | 57.19 ± 1.62 | 60.2 ± 1.95 | 0.23 |
| Female | 23 (0.39) | 22 (0.38) | 0.56 |
| Non-white race | 9 (0.15) | 9 (0.15) | 0.28 |
| **Diagnoses and comorbidities** |  |  |  |
| *C. difficile* infection | 4 (0.07) | 11 (0.18) | 0.07 |
| Leukemia | 21 (0.36) | 17 (0.29) | 0.38 |
| Lymphoma | 12 (0.21) | 10 (0.17) | 0.49 |
| Bone marrow transplant | 15 (0.25) | 14 (0.24) | 0.64 |
| Solid organ malignancy | 35 (0.6) | 40 (0.67) | 0.33 |
| Metastatic malignancy | 29 (0.49) | 35 (0.59) | 0.10 |
| Diabetes | 27 (0.46) | 23 (0.39) | 0.59 |
| Coronary artery disease | 6 (0.11) | 10 (0.17) | 0.72 |
| Congestive heart failure | 19 (0.32) | 18 (0.3) | 0.60 |
| COPD | 21 (0.35) | 36 (0.61) | 0.02 |
| Peripheral vascular disease | 6 (0.1) | 2 (0.03) | 0.31 |
| End stage renal disease | 18 (0.31) | 26 (0.44) | 0.07 |
| Connective tissue disease | 1 (0.01) | 4 (0.06) | 0.35 |
| Peptic ulcer disease | 9 (0.15) | 6 (0.11) | 0.50 |
| Cirrhosis | 2 (0.04) | 9 (0.15) | 0.06 |
| Cerebrovascular disease | 12 (0.21) | 17 (0.29) | 0.54 |
| Hemiplegia | 4 (0.06) | 12 (0.2) | 0.07 |
| Dementia | 1 (0.01) | 3 (0.05) | 0.34 |
| Charlson Score (mean ± SE) | 3.71 ± 0.22 | 4.45 ± 0.25 | 0.05 |

Cases and *controls* were matched by "time at risk" and unit or ward. Values are reported as n (proportion) unless otherwise noted.

### Admission gut microbiota do not predict VRE acquisition

We first compared baseline microbiota across patients who did (*cases*) and did not (*controls*) subsequently acquire VRE. Baseline gut communities of cases and controls did not differ in their community composition, determined either via permutation testing (p=0.30 by PERMANOVA) or via visualization (principal component analysis, **Figure 1a**). Similarly, baseline gut communities of cases and controls did not differ in their species diversity as measured by the Shannon diversity index (mean of 2.72 ± 0.90 for controls, mean of 2.71 ± 0.76 for cases, p=0.96 for all matched case-control pairs) (**Figure 1b**). We concluded that the gut microbiota, as represented by the 16S profile of these samples of hospitalized patients, do not predict subsequent VRE acquisition.

**Figure 1.**
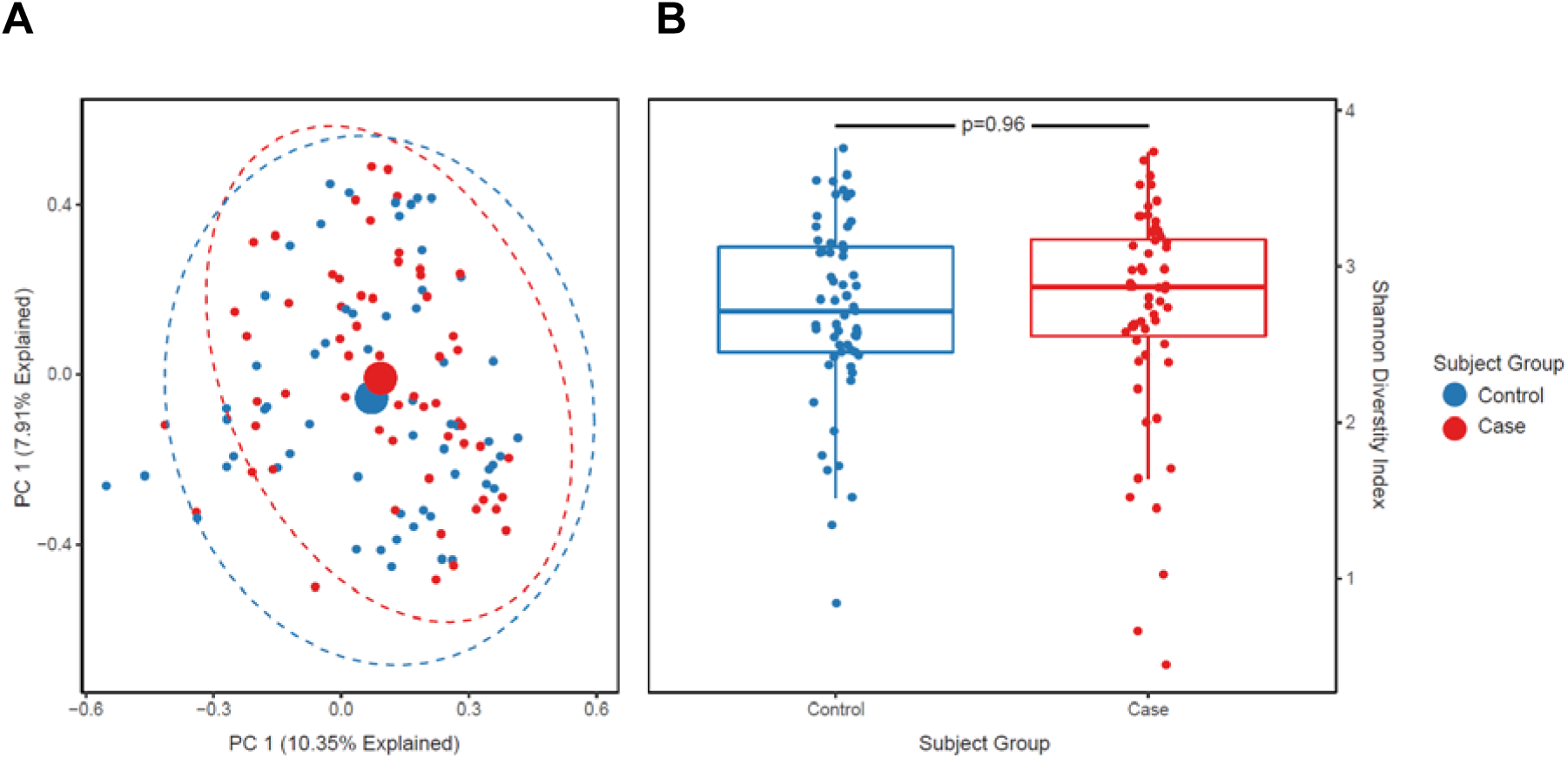
In Hospitalized patients, admission gut microbiota do not predict subsequent VRE acquisition. We used 16S rRNA sequencing to characterize gut bacterial communities in 118 hospitalized patients who tested negative for VRE at admission. We compared admission gut microbiota across 59 matched cases (patients who acquired VRE) and controls (patients who did not acquire VRE). A Visualization of admission gut microbial communities using Principal Component Analysis. The admission gut communities of cases and controls did not differ in their community composition, either visually or via permutation testing (P=0.3, PERMANOVA). B. Comparison of average species diversity as measured by Shannon diversity index in admission gut communities. The admission gut communities of cases and controls did not differ in their community Shannon Diversity index (p=0.96, Conditional logistic regression).

### At the time of VRE detection, the gut communities of cases and controls differ only in the abundance of *Enterococcus*

We next compared gut communities across matched *cases* and *controls* after time-at-risk: after cases had been colonized and time-matched controls had not. After time-at-risk, gut microbiota did differ across *cases* and *controls* (p <0.001, by PERMANOVA), though Shannon diversity index did not (mean of 2.38 ± 0.106 for controls, mean of 2.22 ± 0.115 for cases, p=0.33 for all matched case-control pairs). The difference in gut microbiota was driven by the increased relative abundance of a single OTU, the Enterococcus-classified taxonomic group (OTU0004), which was greater in *cases* than *controls* [p=0.01 via *mvabund*, p<0.001 via Random Forest] (**Figure 2**). When this *Enterococcus* OTU was excluded from the analysis, we found no significant difference in communities across *cases* and *controls* [p=0.12 by PERMANOVA] (**Figure 3**). We thus concluded that at the time of VRE acquisition, the gut microbiota of VRE-infected and uninfected patients differ only in the relative abundance of *Enterococcus*, and do not consistently differ in their *non-Enterococcus* microbiota.

**Figure 2.**
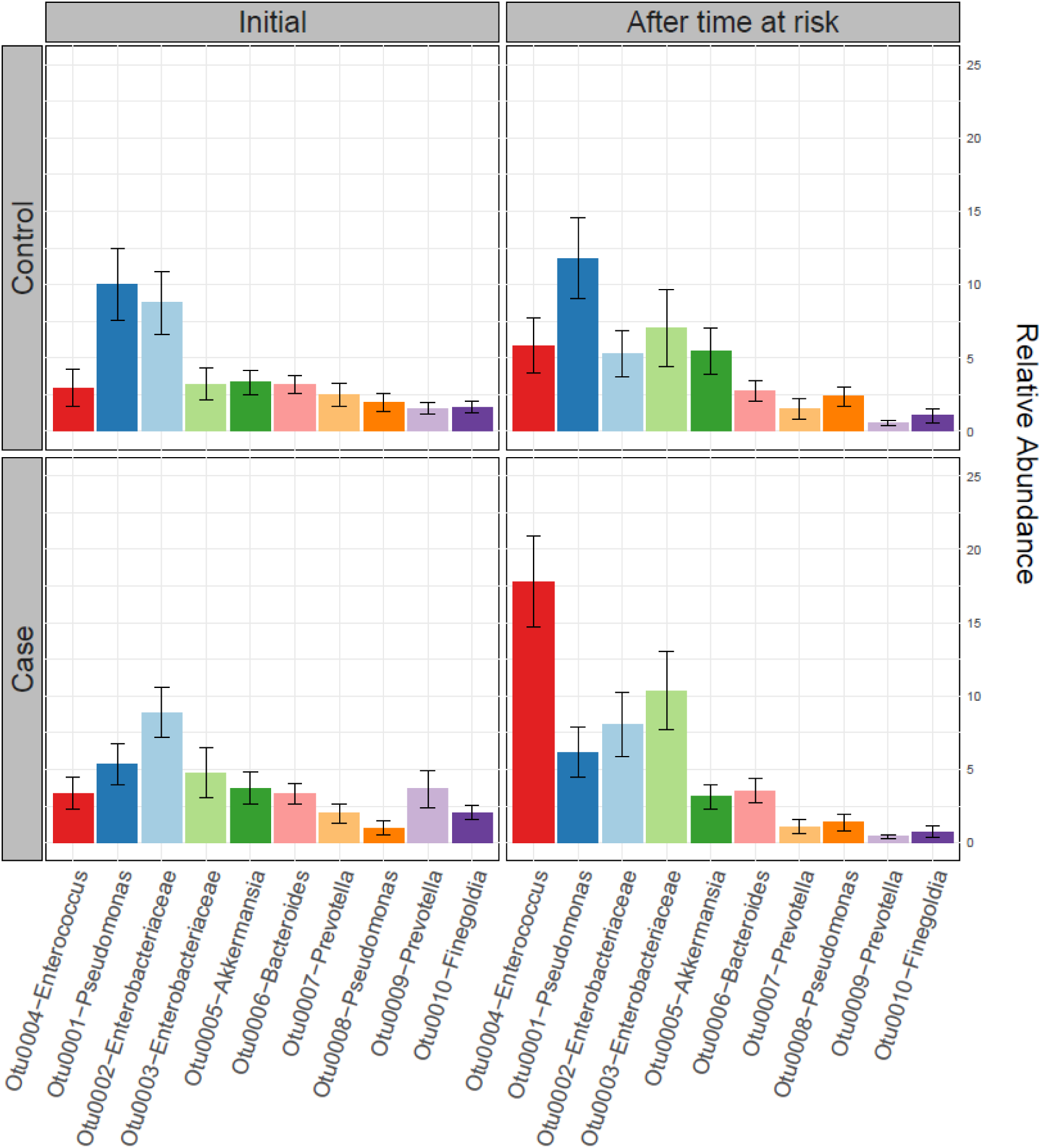
After the time at risk, the gut microbiota of cases and controls differ primarily in their relative abundance of *Enterococcus*. The ten most abundant bacterial taxa are shown in controls (top) and cases (bottom), at the time of admission (left) and following matched time-at-risk (right). Cases and controls did not differ in their admission microbiota (left). After the time-at-risk, the gut microbiota of cases were enriched with *Enterococcus* relative to control (P<0.01, *mvabund*), but otherwise did not differ in their community composition (p >0.05 for all other taxa, *mvabund*).

**Figure 3.**
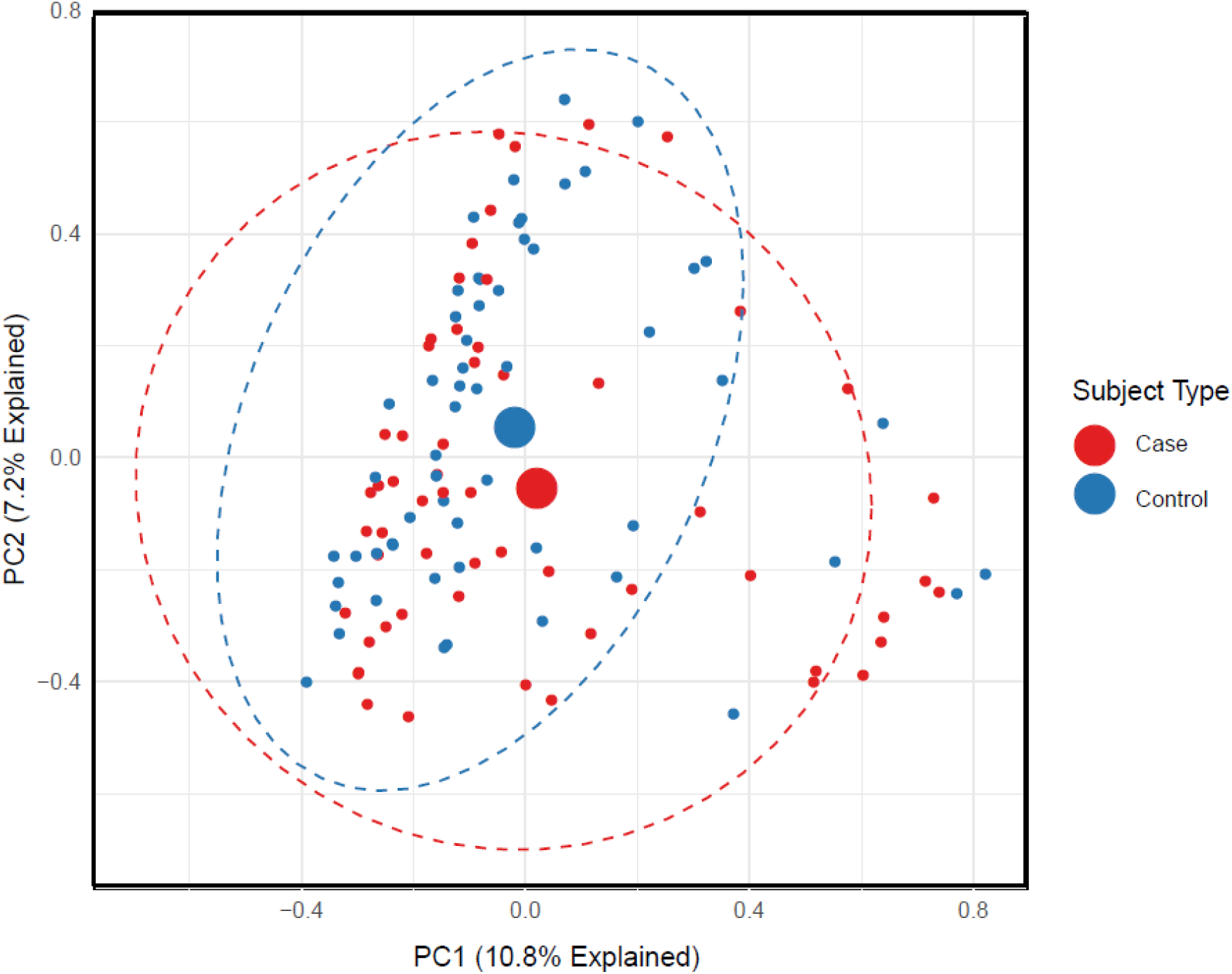
With the exception of *Enterococcus*, gut communities of VRE infected and uninfected patients do not differ. When we excluded *Enterococcus* OTU enriched in VRE-infected patients, we found no remaining difference in bacterial community composition, either visually (Principal Component Analysis) or via permutation testing (P = 0.12 PERMANOVA).

### Gut microbiota change rapidly and profoundly in hospitalized patients

Given the lack of differentiation of gut communities across *cases* and *controls* at admission and at the time of VRE colonization, we then asked if the temporal change in gut microbiota could distinguish the two groups. We did this by calculating the relative dissimilarity of admission and index (time-at-risk) communities for each subject using Jaccard Distance, a metric of dissimilarity between gut microbial communities measured on a scale of 0 (complete similarity) to 1 (complete dissimilarity, **Figure 4**). The gut communities of both groups underwent a rapid, profound change in composition. Within several days of admission, gut communities of both cases and controls bore little similarity to the communities detected at the time of admission. Size of change in gut communities did not differ across cohorts (0.87 ± 0.016 for cases, 0.86 ± 0.017 controls, p=0.84). *Cases* and *controls* also had similar decreases in Shannon diversity (−0.48 ± 0.090 for cases, -0.40± 0.077 for controls, p =0.93 for all matched case-control pairs). We found that Jaccard distance was significantly correlated with time (Spearman’s rank correlation coefficient ρ= 0.32, p =0.0006), and determined that a negative exponential model best fit the data, with gut microbiota approaching complete dissimilarity at an exponential rate of 0.47 * *e*^−0.47*t+32^ (*t* representing the time between swabs). We found no significant difference in the rate of change between the two groups. We noted that the predicted mean Jaccard distance for two rectal swab samples taken on the same day (t=0) was 0.79±0.058, implying a substantial amount of variation in community structure within the same day of admission.

**Figure 4.**
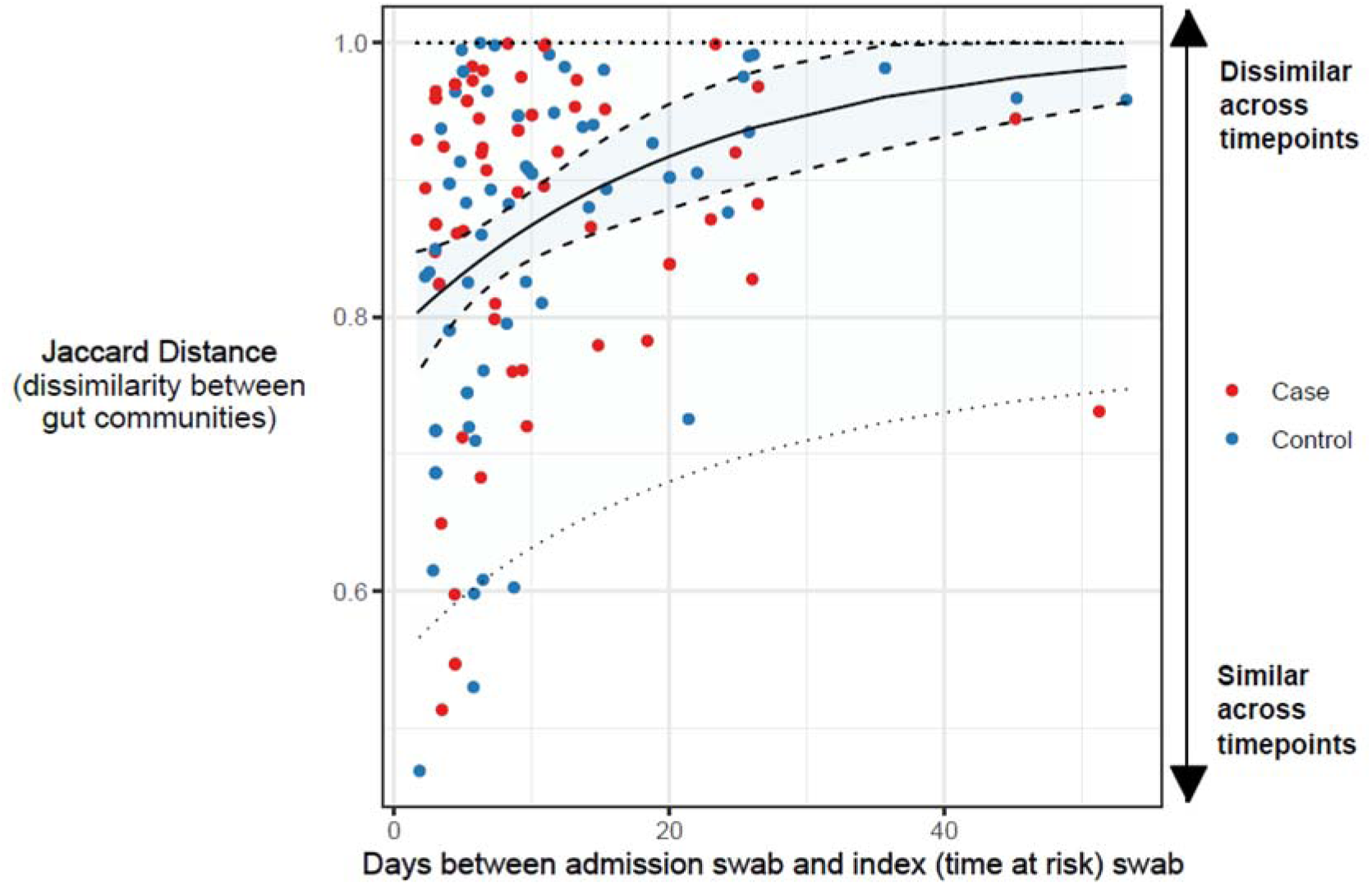
Rapid and dramatic change in gut microbiota among hospitalized patients. We calculated the dissimilarity between admission and subsequent (index, time-at-risk) gut communities in both cohorts with Jaccard distance. Both cases and controls exhibited rapid changes in gut communities during hospitalization, with Jaccard distance changing at an exponential rate. Cases and controls did not differ from each other in temporal disruption of gut microbiota. Dashed lines in the figure represent the 95% confidence interval for predicted mean Jaccard distance (inner ribbon) and predicted Jaccard distance for an individual subject (outer ribbon).

### Gut microbiota on admission are associated with *Enterococcus* expansion

Finding no difference in the community composition, diversity, or temporal rate of change across patients who did (*cases*) and did not (*controls*) acquire VRE during their hospitalization, we asked if gut microbiota on admission could predict the relative *abundance* of *Enterococcus* in VRE colonized patients. We restricted our analysis to case subjects and built a random forest regression model to identify taxa present on admission that were predictive of final *Enteroccus* relative abundance. Only *Blautia* and *Lactobacillus* were significant after correcting for multiple testing and feature importance bias (**Figure 5, Supplemental Table 1, Supplemental Figure 2**). *Blautia* spp (OTU 0092) on admission was predictive of decreased *Enterococcus* (−10.3% relative abundance adjusted p =0.004 by Mann-Whitney U-test), and *Lactobacillus* spp (OTU 0026) was with an increased abundance of *Enterococcus* after the time at risk (+12.5% relative abundance p =0.007 by Mann-Whitney U-Test). Thus, we found that the presence of specific anaerobes previously implicated in *Enterococcus* colonization resistance[2,15,40] are predictive of decreased *Enterococcus* abundance in VRE colonized patients. These findings suggest that VRE *acquisition* and *Enterococcus expansion* are two distinct processes with different risk factors and pathophysiology.

**Figure 5.**
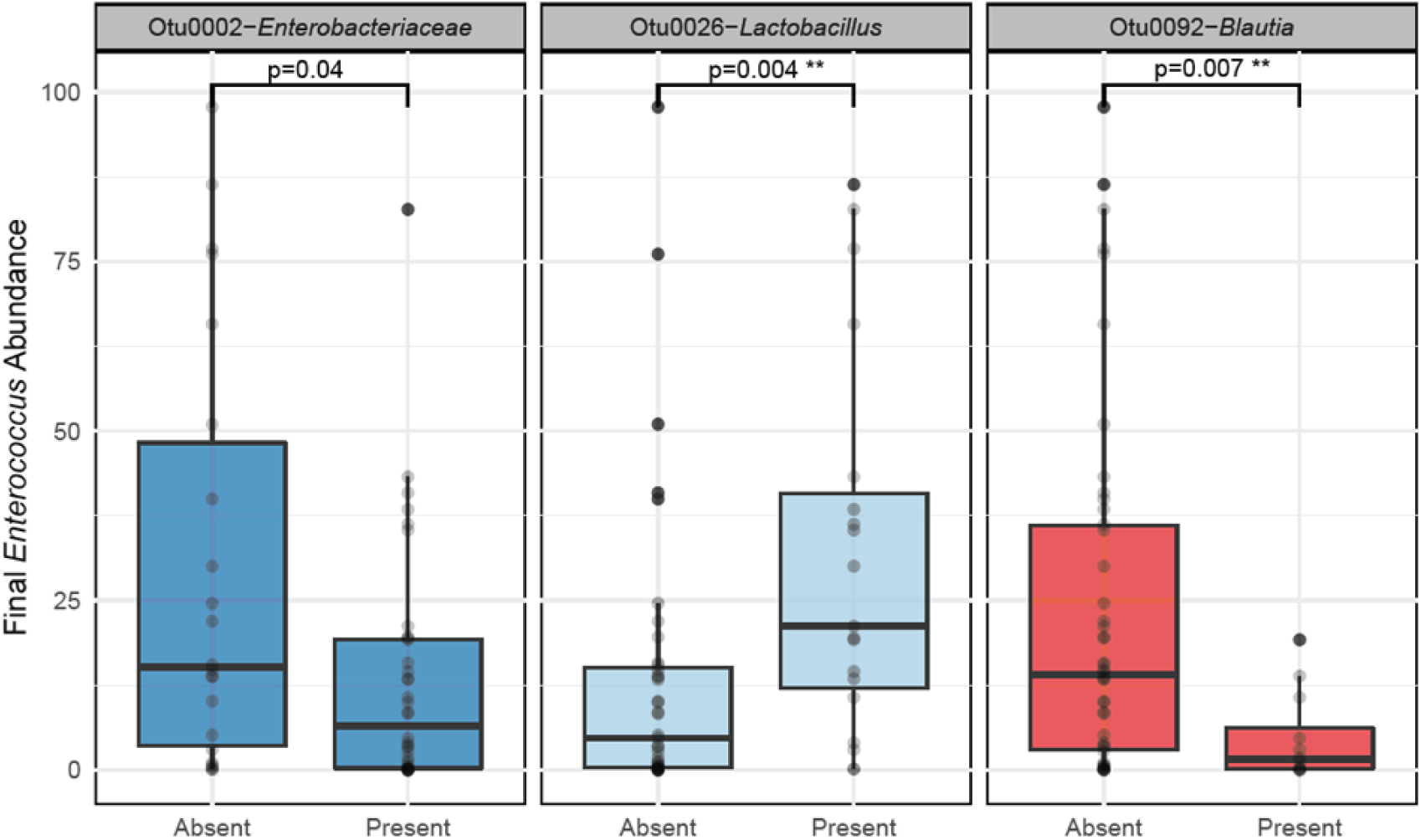
Presence of *Blautia* species on admission is predictive of decreased Enterococcus abundance at the time of VRE acquisition. A random forest regression model identified 7 OTUs present on admission that predicted subsequent relative abundance of *Enterococcus* spp. Of these, the presence of *Enterobacteriaceae* spp., *Lactobacillus* spp., and *Blautia* spp. were significant predictors of final *Enterococcus* spp. relative abundance. Only *Lactobacillus* spp and *Blautia* spp. remained significant after correcting for multiple testing. Significance determined using the Mann-Whitney U test controlled for multiple comparisons.

## DISCUSSION

In this study gut microbiota did not predict VRE acquisition in hospitalized patients. Secondary analysis identified individual members of the gut microbiota that do predict *Enterococcus abundance* at the time of VRE acquisition, implying that *acquisition* and *expansion* of VRE may be distinct processes. The community composition, diversity, and temporal rate of change did not differ across patients who did (*cases*) and did not (*controls*) acquire VRE during their hospitalization. As expected based on the study design, gut communities of cases had a greater abundance of *Enterococcus* than controls after the time at risk.

Gut communities of all subjects demonstrated a rapid and dramatic change during hospitalization that was time-dependent. In this population, antibiotic use was prevalent (Table 2), gut microbial communities were remarkably dynamic (Figure 5), and admission gut microbiota provided very little information about microbiota after the time at risk. Our model of Jaccard distance over time estimated a mean Jaccard distance of 0.79 between two rectal swabs taken on the same day of admission, implying that only 21% of gut microbiota remain constant with resampling within 24 hours. Given the significant correlation between Jaccard distance and time, some of this change is likely due to the disruptive pressures that face gut microbiota upon hospitalization (i.e., antibiotics). However, a large portion of this change may represent stochasticity and noise introduced by variation in sample collection and storage. These results have important implications for the clinical use of gut microbiota for therapy, prediction, and risk stratification. Given the rapid change of gut communities upon hospitalization, a single static 16S analysis of gut microbiota may miss subtle dynamics important for VRE *acquisition* and is subject to a large amount of noise that may obscure a true biologically meaningful association. Future study of the gut microbiota in VRE *acquisition* may need to move beyond traditional 16S analysis, which can be time-consuming and miss important species-level information [41,42]. Real-time metagenomics and rapid, ultrasensitive quantification technologies hold promise as tools with better resolution to evaluate these processes [43,44].

**Table 2.**
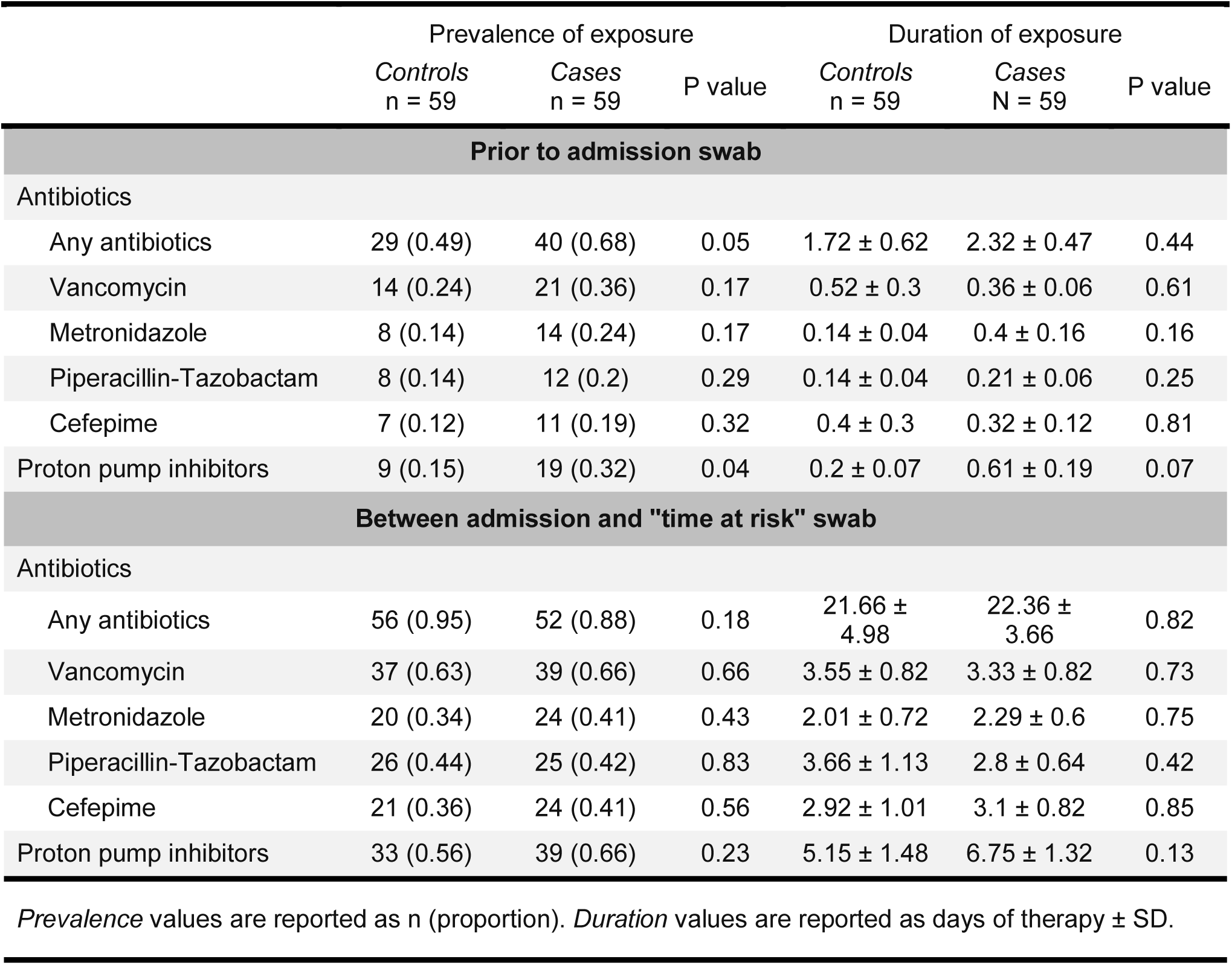
Medication Exposure of Matched Cohorts

Despite the dramatic change in community structure, we did find some evidence of colonization resistance, as admission microbiota *were* predictive of *Enterococcus abundance* at the time of VRE detection. VRE colonized subjects with *Blautia* had less *Enterococcus* expansion, consistent with prior studies [2,15,40]. We hypothesize that there may be a distinction between VRE acquisition and VRE expansion. In conjunction with earlier studies [2,40,45], our findings suggest that commensal anaerobes may play a significant role in suppressing VRE once colonized. In this context, our results further support the possibility of microbiome manipulation to reduce VRE burden even in patients *already colonized* to prevent progression to VRE infection in the individual patient[15,45] or transmission into the surrounding environment and other hospitalized patients[2,10,40].

In this retrospective case-control study, we controlled for multiple confounders with our time and unit matched design. We used machine learning algorithms robust to multi-collinearity and overfitting, and applied permutation heuristics to correct for feature importance bias and decrease our false discovery rate. This study reveals an opportunity for future studies to delineate key differences in pathophysiology between VRE acquisition and domination.

In summary, VRE acquisition and expansion may be two distinct processes, and efforts to manipulate the microbiome to prevent the spread of VRE may be more beneficial in reducing VRE domination in colonized patients than in preventing VRE acquisition in un-colonized patients. Future studies of the role of the gut microbiota in VRE acquisition may need to move beyond single time point 16S analyses and address the role of temporal dynamics and stochasticity of gut microbiota in the acquisition and expansion of VRE.

## ACKNOWLEDGMENTS

The authors would like to thank Aline Penkevich for assistance with acquiring rectal swabs for sequencing.

The authors have no conflicts of interest to disclose.

RC No conflict, CAB No Conflict, KH No Conflict, NF No Conflict, PR No Conflict, RPD No Conflict, RJW No Conflict.

